# Full lockdown policies in Western Europe countries have no evident impacts on the COVID-19 epidemic

**DOI:** 10.1101/2020.04.24.20078717

**Authors:** Thomas Meunier

**Affiliations:** Woods Hole Oceanographic Institution, Falmouth, Massachusetts; Ensenada Center for Scientific Research and Higher Education, Ensenada, BC

**Author notes:** Correspondence to T. Meunier.

## Abstract

This phenomenological study assesses the impacts of full lockdown strategies applied in Italy, France, Spain and United Kingdom, on the slowdown of the 2020 COVID-19 outbreak. Comparing the trajectory of the epidemic before and after the lockdown, we find no evidence of any discontinuity in the growth rate, doubling time, and reproduction number trends. Extrapolating pre-lockdown growth rate trends, we provide estimates of the death toll in the absence of any lockdown policies, and show that these strategies might not have saved any life in western Europe. We also show that neighboring countries applying less restrictive social distancing measures (as opposed to police-enforced home containment) experience a very similar time evolution of the epidemic.

## Introduction

The recent COVID-19 outbreak in Europe has challenged the governments responsiveness in front of an unpredictable and unprecedented situation. Since most countries were unprepared to face such an unexpected epidemic, lack of testing capacities yielded most policies to shift towards social distancing measures rather than modern laboratory-based quarantine [10]. A broad range of public actions were taken in response to the epidemic, from no action at all (Sweden) to full lockdown (Italy, France, Spain and United Kingdom), including police-enforced home containment. Other countries, such as the Netherlands and Germany, opted for a measured response, encouraging social distancing without locking their population down.

While new medical treatments proposed to cure COVID-19 cases are required to be validated through controlled double blind studies, the benefits and risks of social distancing strategies are not subject to any comparative tests. However, full lockdown measures, such as those decided in Italy, France, Spain and United Kingdom have not been experienced in Western Europe countries for centuries, and their effects in contemporary population’s mental and physical health is largely unknown. The COVID-19 epidemic episode was shown to, by itself, affect mental health, including anxiety syndromes and depression [21] and the consequences of isolation could enhance these conditions. In the absence of any control group, the impacts on western Europe’s population will not be measurable until months. Nevertheless, increased mortality due to difficulties of access to basic health care, increased mental conditions linked to isolation, as well as social consequences of economic recession, despite being unquantifiable so far, is to be expected. Such measures are thus only appropriate if their impacts on limiting the epidemic spreading save more lives than their inherent death toll. Attempting a real-time assessment of full lockdown policies efficiency thus seems crucial to help public action decisions in the forthcoming weeks.

Recent modeling results suggestes that China’s full lockdown policy was successful in containing the epidemic [9]. In an attempt to predict the efficiency of similar policies in Western Europe countries (Italy, France, Spain and United Kingdom), Picchioti el al. (2020) [16] implemented a SEIR model, testing different lockdown parameterizations, and suggested that early public containment measures could be efficient. However, as acknowledged by the authors, real-time parameterization of a model for an unknown disease is a difficult and uncertain task, and the effects of lockdown may vary from one country to another. Although modeling studies offer valuable insights and possible scenarii for forthcoming events, and might provide a deep understanding of the epidemic’s dynamics *a posteriori*, they require validation, which can only be provided by thorough data analysis. In that regard, the observational efforts of Tobias (2020) [20] represent an interesting approach. The latter recently claimed that the full lockdown policies in Spain and Italy have had positive results in slowing the epidemic. However, their methods, based on fitting linear trends to the logarithm of the daily new cases and daily death numbers, and comparing them before, and after the lockdown policies, might not be appropriate. As will be shown below, to assess the trajectory of the epidemic, one should look for trends in the time derivative of the logarithm of daily numbers, rather than trends in the logarithm of the daily numbers itself.

Here, we show that the available data exhibit no evidence for any effects of the full lockdown policies applied in Italy, Spain, France and United Kingdom in the time evolution of the COVID-19 epidemic. Using a phenomenological approach, we compare the evolution of the epidemic before and after the full lockdown measures are expected to produce visible results. Our approach have similarities with Tobias (2020)’s [20]: it is focused on incident rather than cumulative data, and it compares pre-lockdown and post-lockdown trends. However, here, no positive changes are noticed in the trend of the daily death growth rate, doubling time, or reproduction number, weeks after lockdown policies should have impacts.

## 1 Methods

Although Epidemic outbreaks are complex dynamical systems, the daily new cases number most generally follows a similar time evolution: after an exponential growth, infections slow and eventually decay exponentially as, whether group immunity is reached, or seasonal factors or public actions slow the virus reproduction. This behavior has been observed for seasonal influenza [14], H1N1 [7] as well as for recent coronavirus epidemics such as SARS [1, 5, 8] or MERS [3]. It is usually well described by exponential functions such as the logistic distribution or the Gauss function. The Gauss function is defined here for time evolution of new cases as:

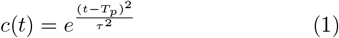

where *c*(*t*) is the daily number of new cases, *t* is time, *T*_*p*_ is the time of the peak (maximum infection), and *τ* is a time scale defining the duration of the epidemic. In the assumption of a steady relationship between the number of cases and the number of fatalities (the fatality rate *µ* is time-independent), the daily death number *d*(*t*) is linked to the daily new cases number *c*(*t*) through the time-lagged proportionality relation:

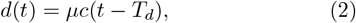

where *T*_*d*_ is the average time between infection and death. It thus follows a similar Gaussian-like law. However, the latter never is a purely Gaussian function, and is often skewed or exhibits more complex patterns [8], so that the definition of Eq (1) is not exactly correct and needs to be generalized.

It is natural and convenient to express time evolution of *c*(*t*) as a power function with a time varying exponent. As for any strictly positive function, *c*(*t*) can be written in the form of a generalized exponential function:

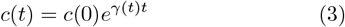

where the time varying function *γ*(*t*) will be referred to as the growth rate of *c*(*t*). Expressing time evolution of the daily new cases number in the form of Eq (3) can be thought of as a generalization of Richards’s phenomenological model [18].

If *γ* is a constant, *c*(*t*) is a pure exponential. If *γ*(*t*) is a linearly decaying function of the form *γ*(*t*) = *β* + *αt*, with *α <* 0, *c*(*t*) is a Gauss function, and *T*_*p*_ and *τ* can be expressed in terms of the slope and intercept of *γ*(*t*):

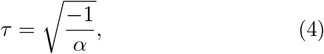

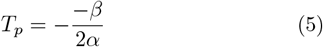

In that case, the short time asymptotic limit is close to a pure exponential growth, since *t << T*_*p*_. It then slows down as *t* approaches *T*_*p*_, and starts to decay as *t* passes *T*_*p*_. Eq (3) however allows any form of *γ*(*t*) and is not restricted to Gaussian or exponential behaviors. Values of *γ*(*t*) can be retrieved from any time series of the daily new cases number, or equivalently the daily death number :

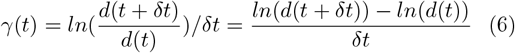

*γ*(*t*) can thus be defined as the time derivative of the natural logarithm of the daily death number. This method is commonly used in the study of transient perturbations growth in fluid mechanic’s generalized stability theory [6, 12]. In this work, we will be primarily studying the time variations of *γ*, and search for visible trends in the latter.

Since the time necessary for the number of fatalities to double (hereafter doubling time) is a commonly used diagnostic of an epidemic evolution, it is also computed in this work. The total number of deaths at time t is the time integral of the daily death number:

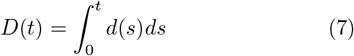

Since *D*(*t*) is also a strictly positive function, it can be expressed in a similar form as Eq (3):

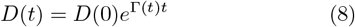

and Γ(*t*) can be retrieved as in Eq (6):

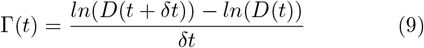

The doubling time (*T*_2_) is related to Γ following:

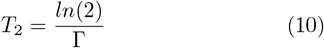

so that we can obtain instantaneous estimates of *T*_2_(*t*) without waiting for the number of total fatalities to actually double.

To assess the efficiency of lockdown policies, we first compute the growth rate *γ*(*t*) from the daily deaths observations and apply linear regression to estimate its trend before the lockdown should have any visible effects (*t < T*_*ld*_ + *T*_*d*_, where *T*_*ld*_ is the start date of the lockdown measures). We then predict values for each variable of interest after the lockdown should have visible effects by extrapolating the linear evolution of *γ*(*t*) after this date. This allows us to compare observed values of growth rate, daily deaths, doubling time, and total fatalities number, with the values expected from the pre-lockdown trend (what would have happened if nothing had changed).

To assess the evolution of the epidemic with a more classical approach, we also compute an instantaneous reproduction number as well as an estimate of the reproduction number, based on the daily deaths data and Eq (2), which links the daily deaths number, the fatality rate, and the daily new cases number. The reproduction number is the number of secondary infections provoked by a typical case [1]. In practice, the reproduction number shows large variability depending on a number of factors such as the age or the region [15]. However, a mean estimate is useful to assess the epidemic stage. Here, we approximate the instantaneous reproduction number *R*_*i*_ as the ratio of the number of new cases and the total number of contagious cases at time *t*:

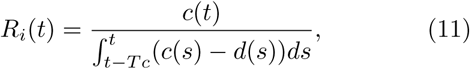

where *T*_*c*_ is the time during which an infected person remains contagious, and *s* is an integration variable. It is important to note that we make the assumptions that new cases start to be contagious right after infection (zero generation time), and that all cases within the contagion period *T*_*d*_ are equally likely to produce secondary infections. While our simplifying hypothesis of zero generation time might yield to an underestimate of the reproduction number, it does not affect its general trend, which is what this study is focused on. The reproduction number *R*, which is the number of persons that will be infected by each contagious person during the time *T*_*c*_ is approximated as:

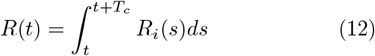

The epidemic is in a growing phase if *R >* 1 and decays otherwise.

## 2 Data

Because of the important proportion of asymptomatic cases of COVID-19 [2, 17, 13] and of the testing policies of most countries, which are restrained to severe and potentially deadly cases, the daily number of new confirmed cases is not a reliable variable to assess the evolution of the epidemic. We thus only used the daily deaths number to estimate the growth rates and their trends. The daily number of new cases is inferred from the daily death number and a fixed fatality rate using Eq (2).

The data used in this study are produced by the European Center for Disease Prevention and Control (ECDC). Because of a lack of daily deaths reports in nursing homes in France until 02 April 2020, that were suddenly corrected in the dataset, we only use hospital deaths data for this country, for consistency of the time series. The daily and total deaths numbers in France are thus greatly underestimated in our study, but one should keep in mind that we are focused on time evolution and trends rather than absolute values, so that time-consistency is the single most important requirement for the data.

To compute the growth rate of the daily deaths number *γ*(*t*) and of the total fatalities number Γ(*t*) as well as the doubling time *T*_2_(*t*), the daily deaths data are first low-pass filtered using a 3-days running mean on the logarithm *ln*(*d*(*t*)).

The values used here for the time between infection and death *T*_*d*_ ranges between 14 and 20 days, with a median reference value of 17 days. It corresponds to the averaged value of hospitalization to death reported by Russell et al. (2020) [19] (13 days) plus a period of 1 to 7 days between infection and hospitalization. Two different values of the time during which an infected person remains contagious (*T*_*c*_) were tested to infer the reproduction number: 14, and 21 days. The former is the duration of the quarantine applied to any confirmed cases in most countries, and the latter is a longer estimate used for comparison since the 14 days value is uncertain [19]. For computing estimates of the daily case number, we used a fatality rate of 1.7 %, which is a median value between Russell et al. (2020)’s [19] estimates of the Infection Fatality Ratio and Case Fatality Ratio onboard the Diamond Princess passenger ship. The latter also closely matches South Korea’s fatality rate (1.6%) [4], which is one of the most reliable national estimate so far, given the wide-range testing policy and the advanced stage of the epidemic in this country.

## 3 Results

Time evolution of the reproduction numbers *R*_*i*_ and *R* is shown in figure 1 for France, Italy, Spain, and United Kingdom. 4 different estimates are proposed in each figure: the estimates defined in Eq (12), computed for values of contagion duration of *T*_*c*_ = 14 and 21 days, as well as values of the instantaneous reproduction number multiplied by the contagion duration (*R*_*i*_*T*_*c*_). For all four variables, time evolution exhibits a similar pattern: a steady decreasing trend from 3.5 to 6 secondary infections per case in the beginning of the epidemic to less than unity 20 to 40 days before the reference date (April 24). In all four countries, no discontinuity in the general decaying trend is observed around the full lockdown’s start date. Even though this date coincides approximately with the 1-crossing of *R* in France, Italy, and Spain, the latter is only the follow-up of a longer term decay. Note that, despite the simplifying assumptions used here, our estimates are of the same order of magnitude as Liu et al. (2020)[11]’s interval [1.4-6.49] for the outbreak in Wuhan.

**Figure 1:**
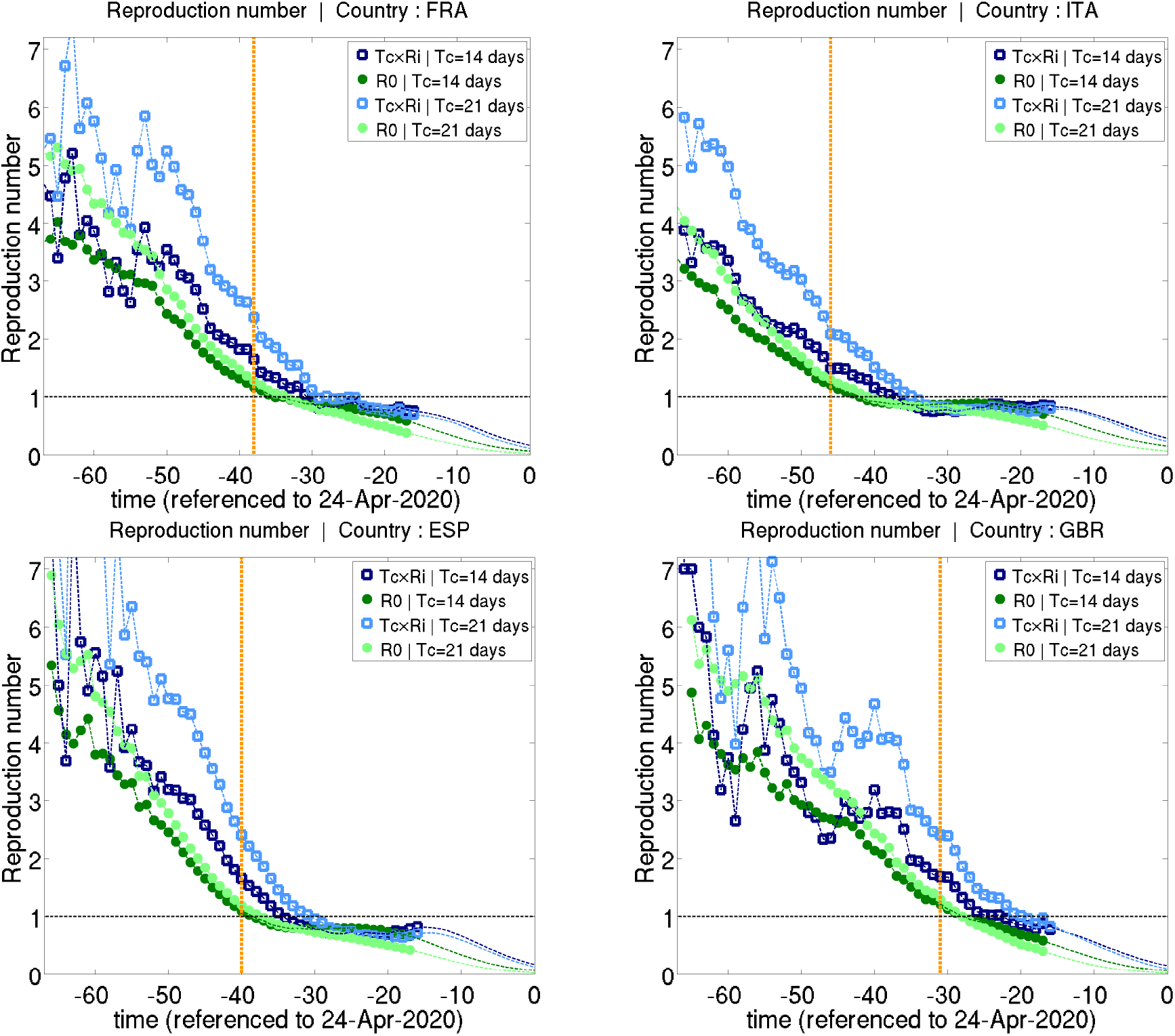
Time evolution of the reproduction number for France, Italy, Spain, and Great Britain. The blue square represent the daily reproduction number multiplied by an estimate of the time during which a case is contagious (*R*_*i*_*T*_*c*_). The green dots represent the reproduction number (*R*) computed as a time integral of the daily reproduction number (*R*_*i*_) and thus takes into account time variations of the latter. The dashed vertical orange line is the start date of the lockdown policies. Two values of *T*_*c*_ were used for each variable: 14 and 21 days. Time is referenced to April 24, 2020.

Analysis of the evolution of the growth rate *γ*(*t*) confirms this long-term trend in the trajectory of the epidemic before any full lockdown policies were effective (figure 2). A general decaying trend of *γ*(*t*) is evident from the beginning of the epidemic in all 4 countries, although some variability exists around the linear trend, with a nearly periodic oscillation of 5 to 8 days. Linear regression satisfyingly models the time evolution of *γ*, with coefficients of determination *r*^2^ (fraction of the variance explained by the model) of 0.67, 0.72, 0.73, and 0.61 for France, Italy, Spain, and United Kingdom, respectively. Linearity of *γ*(*t*) suggests that time evolution of the epidemic is consistent with a Gauss function. Comparing the linear decaying trend before, and after the time by which full lockdown policies should have visible impacts, we find that the slope of *γ*(*t*) decreases in France, Italy, and Spain after the full lockdown, and remains constant in United Kingdom. The decay of the epidemic has thus slowed since the lockdown is effective. Comparison of the general decay trends in France, Italy, Spain and United Kingdom with that of the Netherlands provides a further insight on the effects of full lockdown: Netherlands decay trend is slightly slower than France and Spain before lockdown, and is nearly similar to Italy and United Kingdom’s. In all four countries, the decay trend after the effective lockdown is slower than Netherlands trend.

**Figure 2:**
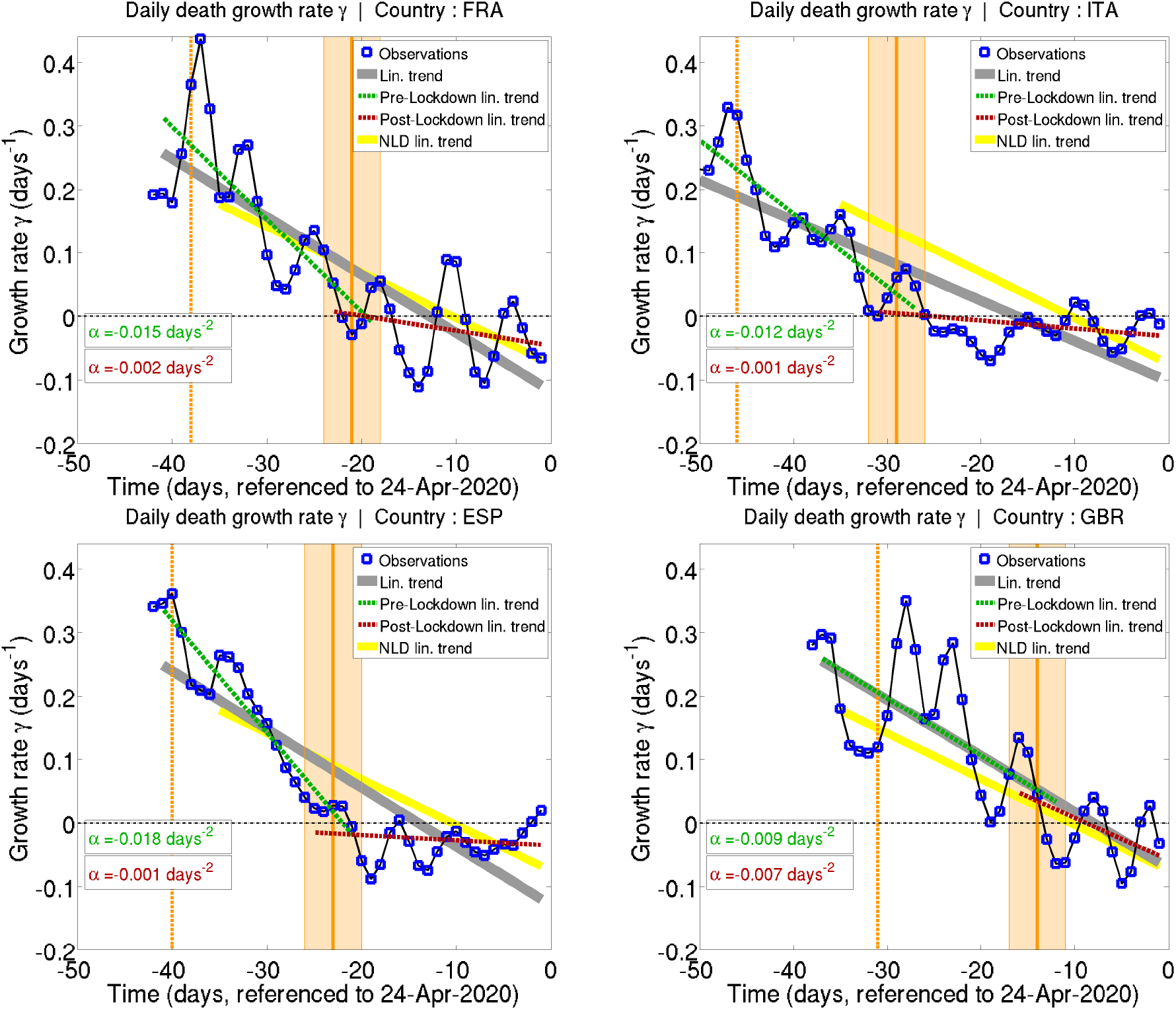
Time evolution of the growth rate of the daily deaths number (*γ*(*t*)) for France, Italy, Spain, and Great Britain. The vertical orange dashed line shows the start date of the full lockdown policies. The orange shaded area represents the time at which the lockdown should show some effects in the epidemic spreading (14 to 20 days), and the thick line is the reference date (17 days). The blue squares represent the observations, and the thick gray line represents the linear trend of the observations. The dashed green and red lines represent the linear trends before and after the lockdown should affect the observations. Time is referenced to April 24, 2020.

Since a raw, visual analysis of the effects of full lockdown on the doubling time could mislead to the impression that its increase is accelerated after the lockdown is effective, we should carefully inspect the results of figure 3. We computed an estimate of the doubling time, assuming the pre-lockdown trend in *γ*(*t*) remains constant after the lockdown is effective (we assume that *γ*(*t*) keeps on linearly decaying after lockdown with the same slope as before lockdown). Comparing these estimates (dashed green line) with the observed values (blue squares) shows that the initial pre-lockdown trend yields a steeper rise in the doubling time than what is observed after lockdown policies should have visible impacts. The visual impression of an accelerating *T*_2_ growth in the data is thus not to be attributed to the lockdown effects, but rather to the inherent growth of the *T*_2_ function when Γ(*t*) reaches small values. Figure 3 thus also underlines the lack of evidence of any effects of the full lockdown.

**Figure 3:**
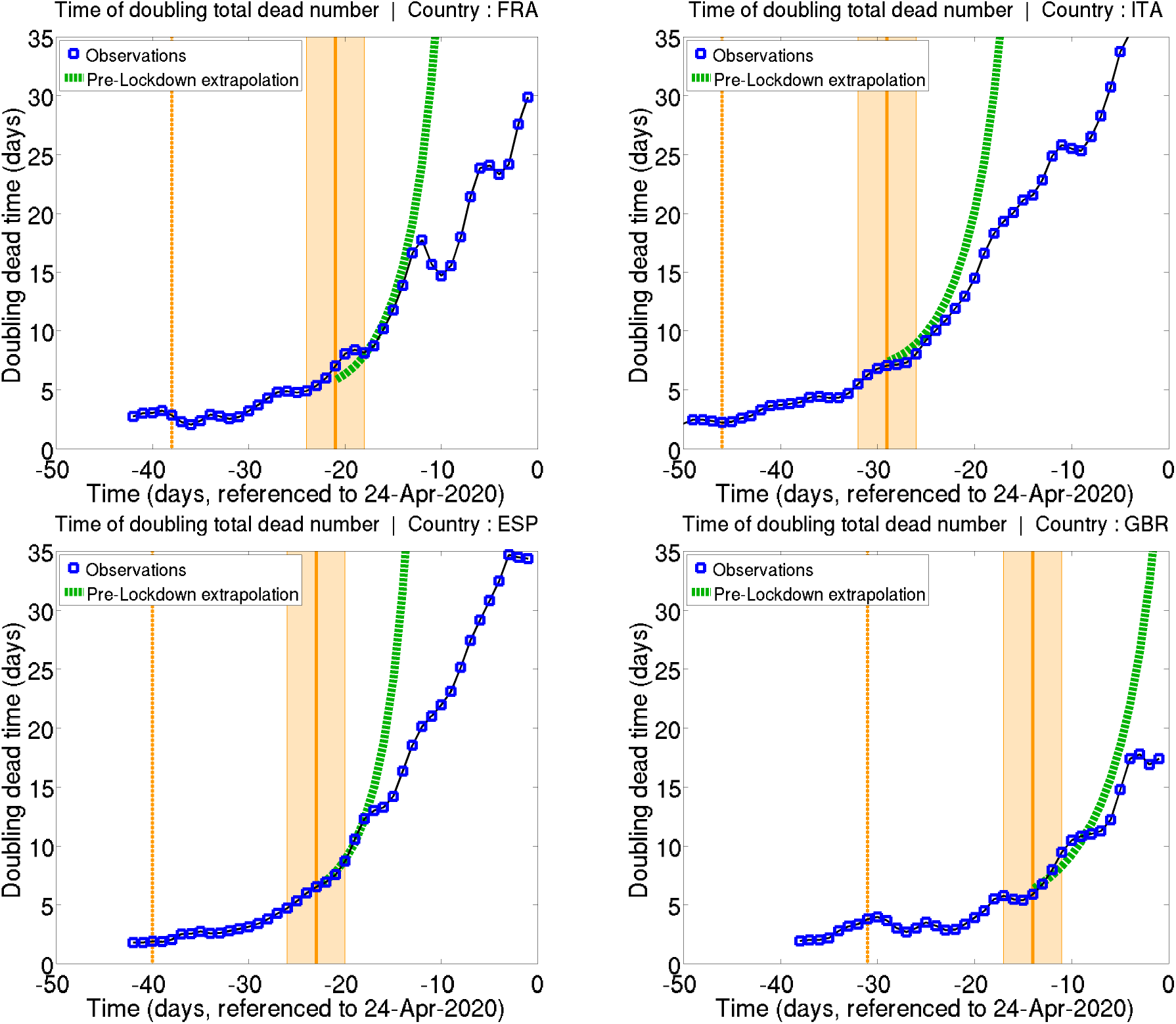
Same as figure 2 for the doubling time. The thick dotted green line represents the expected evolution if the pre-lockdown linear trend in *γ*(*t*) is extrapolated beyond the day lockdown policies are expected to be effective (What would happen without any lockdown, assuming that the growth rate’s evolution remains the same). Time is referenced to April 24, 2020.

**Figure 4:**
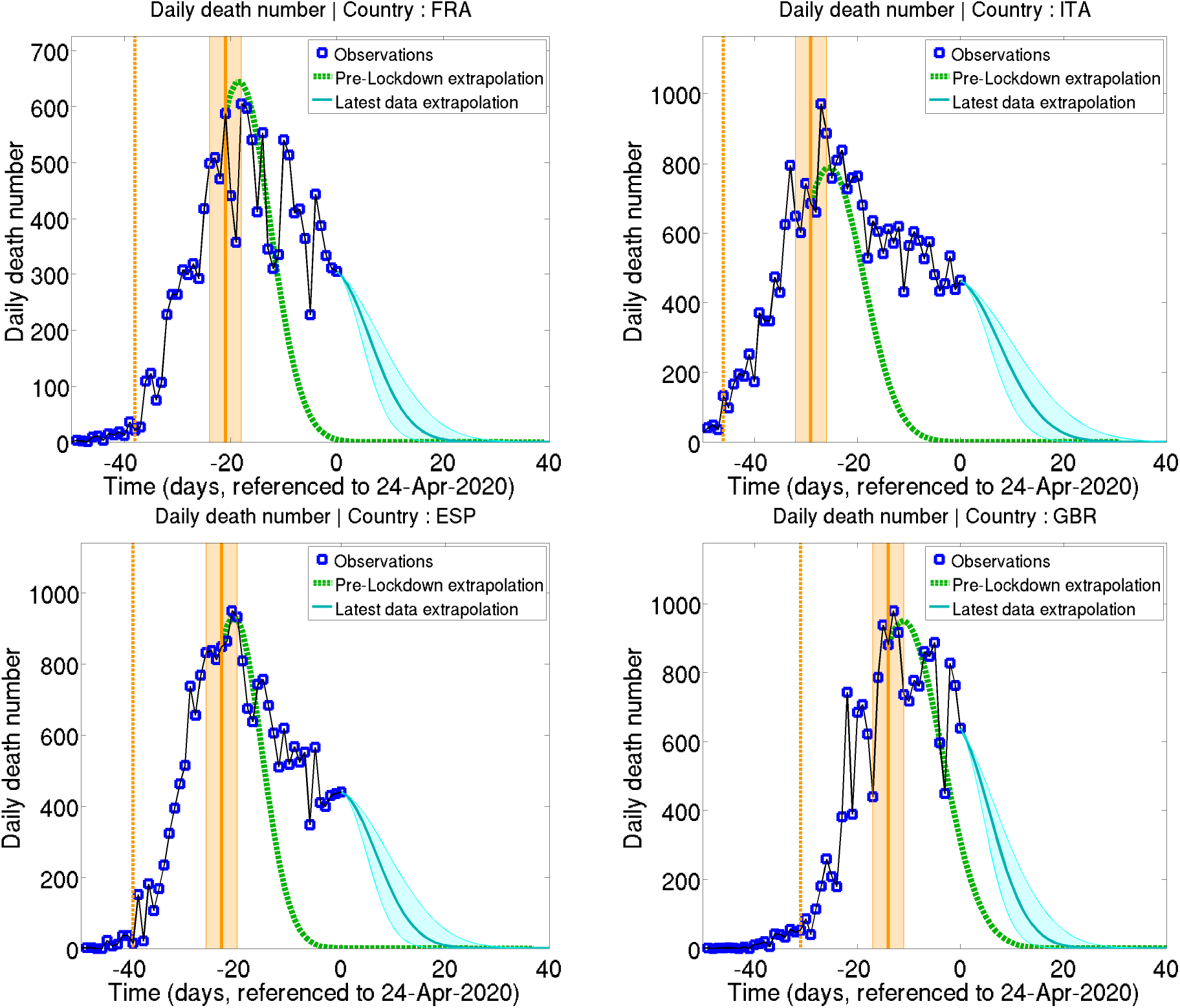
Same as figure 3 for the daily deaths number. The green dotted lines represents the expected evolution based on the pre-lockdown trend of *γ*(*t*), while the plain blue line represents the expected future evolution based on the full time series trend of *γ*(*t*). The light blue shading represents an error margin computed by doubling or dividing by two the slope of the linear fit to *γ*(*t*). Time is referenced to April 24, 2020.

Similarly to the doubling time, we estimated the evolution of the daily deaths number in the hypothesis of a continuation of the pre-lockdown trend in *γ*(*t*) after the lockdown policies should have visible impacts. Our results show that, even though the dates of the daily deaths peaks in France, Italy, and Spain roughly correspond to the dates where lockdown effects should be visible, the peak dates expected from the pre-lockdown trends are actually the same. Moreover, daily deaths observations after this date show a slower decay than what would be expected from the pre-lockdown tendencies. Forecast of the future evolution of the daily deaths number using the same method with the linear trend of the full time series and the latest observations is also shown as the blue line (for indicative purpose only).

Time evolution of the total death toll, both observed, and predicted from pre-lockdown trends is shown in figure 5. One would expect total dead numbers to rapidly saturate at a value close from that corresponding to the crossing of the curve and the date of expected visible lockdown effects. However, the total dead number kept on growing after this date, closely following the values expected from pre-lockdown trends, and even reaching values beyond the death toll expected from the latter. Again, the forecast dead number obtained from extrapolating *γ*(*t*)’s linear trend in the future is presented for indicative purpose. It is however interesting to notice the consistency in the order of magnitude of the final total deaths forecast at a 15 to 30 days interval (at the time of the lockdowns and on April 24.).

**Figure 5:**
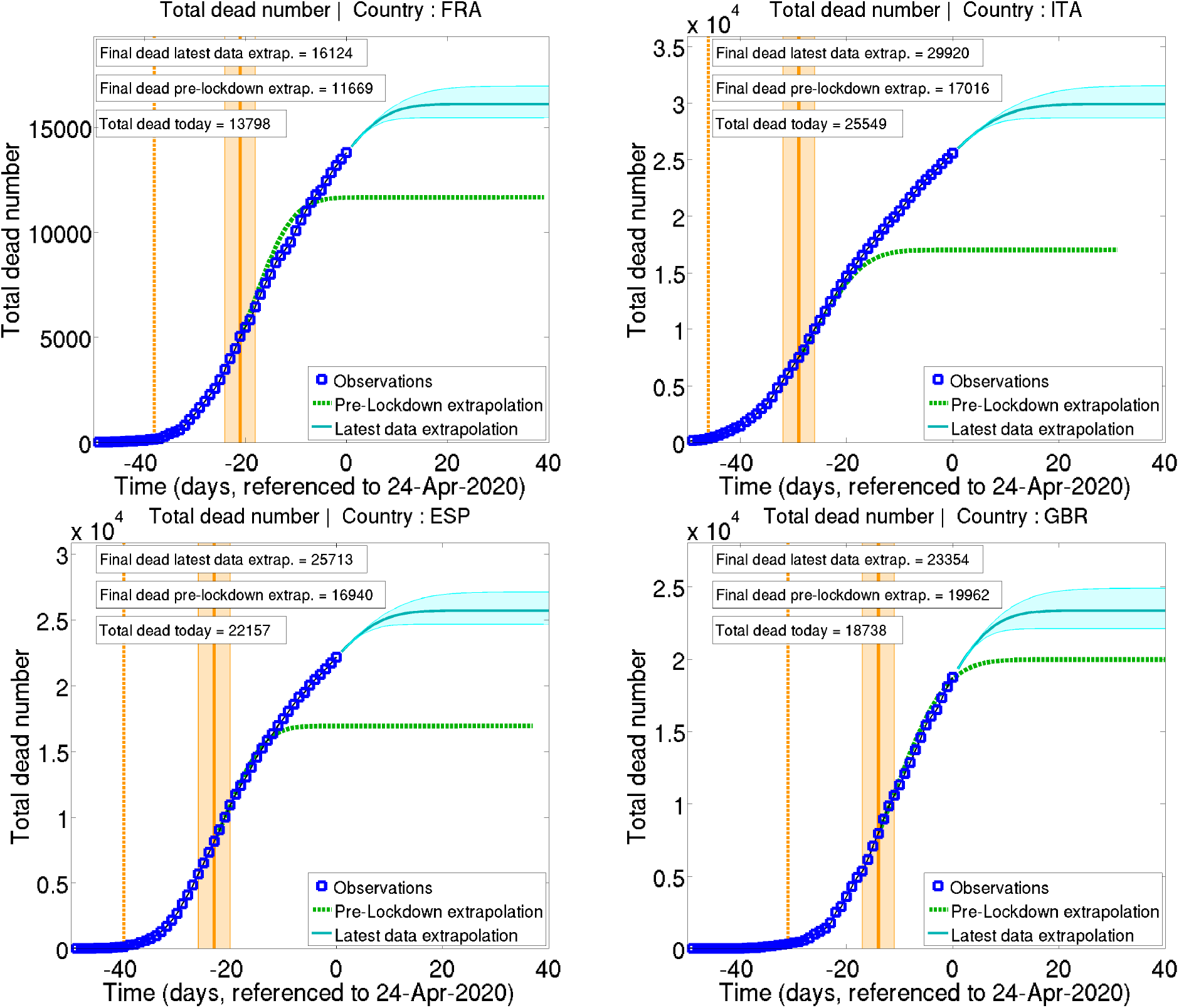
Same as figure 4 for the total fatalities number.

While comparing pre and post lockdown time evolutions of the epidemic brings a useful insight on the impacts of home containment, it is also of interest to compare this evolution with that of countries applying different policies. Figure 6 shows the evolution of the daily deaths growth rates *γ*(*t*), their linear trends, the reproduction numbers, and the doubling times for 10 countries. We selected countries that had over 1000 fatalities by April 15, 2020, and chose to exclude data from China, given the growing doubts on their accuracy. The time reference was chosen to be the day by which the total death toll exceeded 100 in each country. Evolution of *γ*(*t*) shows a similar general decay trend in all countries. It is interesting to note that, while the linear trends of the growth rates have similar slopes in nearly all countries, they show a wide range of intercepts (value of *γ* at t = 0), showing that although the slowdown of the epidemic follows a similar trajectory, each country started at very different levels of growth rate. This general decay trend is accompanied by a regular decay in the reproduction number in all countries, with similar slopes and, again, a wide range of initial reproduction numbers. As expected from a decreasing growth rate and reproduction number, the doubling time is increasing in all countries from the beginning of the time series. Figure 6 thus shows that time evolution of the epidemic is homogeneous in Western Europe, and that the main differences reside in the initial conditions at the beginning of the epidemic. In particular, the figure shows that countries with social distancing policies, but no home containment, such as the Netherlands and Germany experience a very similar decay of the epidemic in terms of growth rate, reproduction number, and doubling time, to countries with police-enforced home containment. On the other hand, results for Sweden suggest that taking no action at all may yield a more variable decay of the epidemic.

**Figure 6:**
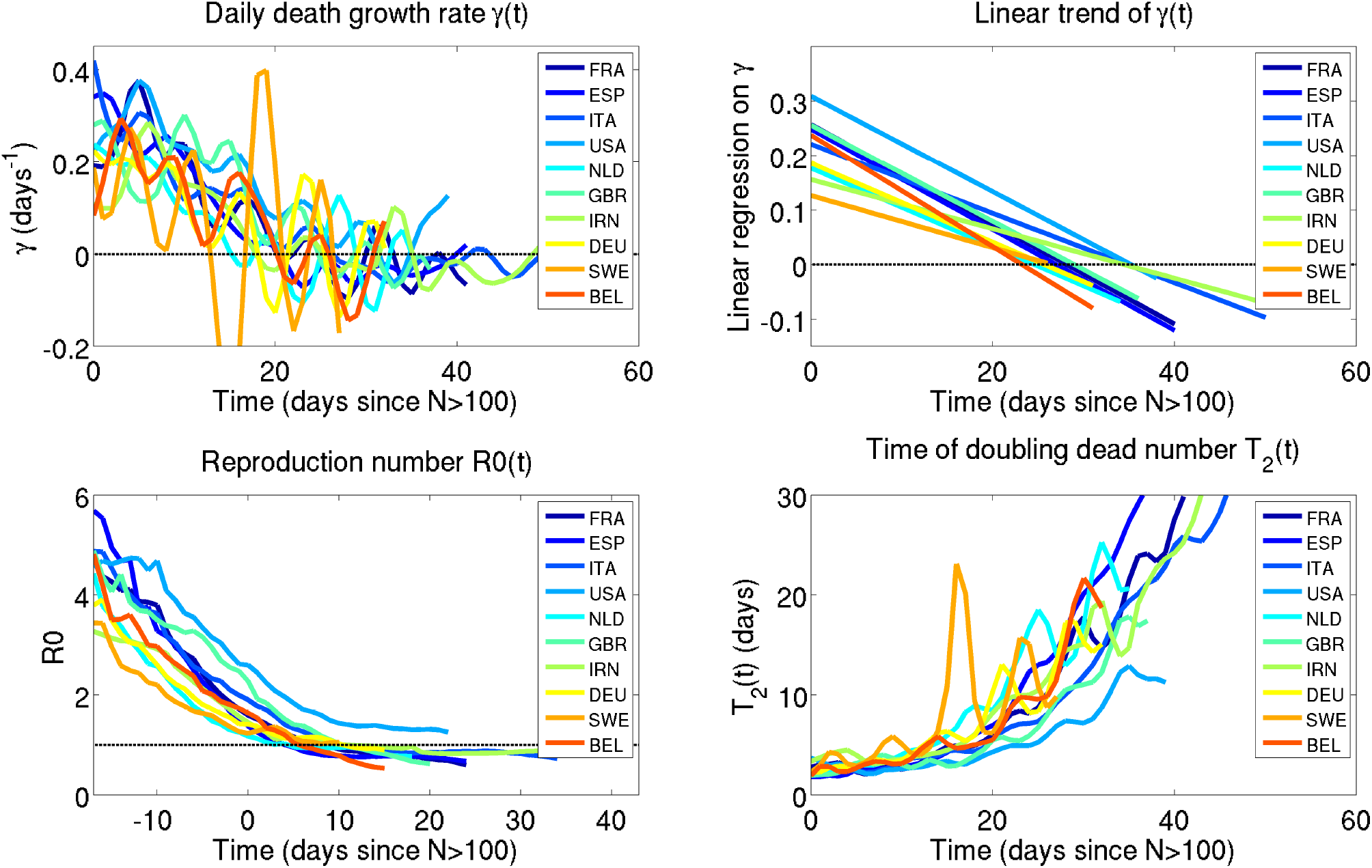
Comparison of the time evolution of the epidemic for 10 countries. The time reference is chosen to be the day each country reaches a total dead number of 100. The top left hand side panel shows the time evolution of *γ*(*t*); the top right hand side panel shows the linear fits to *γ*(*t*); the bottom left hand side panel shows values of the reproduction number *R*(*t*); the bottom right-hand side panel shows the instantaneous doubling time. Time is referenced to April 24, 2020.

## 4 Summary and discussion

This observational study, using a generalized phenomenological method based on official daily deaths records only, shows that full lockdown policies of France, Italy, Spain and United Kingdom haven’t had the expected effects in the evolution of the COVID-19 epidemic. Our results show a general decay trend in the growth rates and reproduction numbers two to three weeks before the full lockdown policies would be expected to have visible effects. Comparison of pre and post lockdown observations reveals a counter-intuitive slowdown in the decay of the epidemic after lockdown. Estimates of daily and total deaths numbers using pre-lockdown trends suggest that no lives were saved by this strategy, in comparison with pre-lockdown, less restrictive, social distancing policies. Comparison of the epidemic’s evolution between the fully locked down countries and neighboring countries applying social distancing measures only, confirms the absence of any effects of home containment. Evolution of the epidemic in Sweden however indicates that, in the absence of any social distancing measures, the epidemic’s decay may be subject to larger fluctuations. This work thus suggests that social distancing measures, such as those applied in the Netherlands and Germany, or in Italy, France, Spain, and United Kingdom before the full lockdown strategies, have approximately the same effects as police-enforced home containment policies.

So far, the reasons for the relatively regular decay of the epidemic remain largely unknown. While social distancing efforts may contribute to it, environmental conditions could as well have played a role (possible seasonality of the virus). The group immunity hypothesis, though being unlikely if the reference fatality rates are correct, deserves a short discussion: computing the number of daily new cases from the number of daily deaths following Eq (2), and using a fatality rate of 1.7%, we forecast a ratio of infected population at the end of the epidemic of 1.4%, 3.0%, 3.2%, and 2.1% in France, Italy, Spain, and United Kingdom, respectively. The latter is obviously far from being able to yield any group immunity. Under the rough assumption that 50 to 70% of the population needs to be infected to ensure group immunity, it is possible to compute an hypothetical fatality rate using Eq (2). We find that, if group immunity was responsible for the decay of the epidemic, the fatality rates would be of about 0.05%, 0.10%, 0.11%, and 0.07% for the 50% hypothesis in France, Italy, Spain, and United Kingdom, respectively, and of 0.03%, 0.07%, 0.08%, and 0.05% for the 70% hypothesis. Obviously, this is only a gedanken experiment, which is far beyond the scope of this paper, and only serological tests and further data analysis, once the epidemic is completely instinct, will allow to discriminate between the possible reasons for its decay.

As a concluding remark, it should be pointed out that, since the full lockdown strategies are shown to have no impact on the epidemic’s slowdown, one should consider their potentially high inherent death toll as a net loss of human lives.

## Data Availability

All data used in this manuscript are publicly available, and all sources are provided.

https://www.data.gouv.fr/fr/datasets/donnees-hospitalieres-relatives-a-lepidemie-de-covid-19/

https://coronavirus.jhu.edu/us-map

